# Protocol for an automated, pragmatic, embedded, adaptive randomised controlled trial: behavioural economics-informed mobile phone-based reminder messages to improve clinic attendance in a Botswanan schools-based vision screening programme

**DOI:** 10.1101/2022.03.21.22272266

**Authors:** Luke Allen, Bakgaki Ratshaa, David Macleod, Nigel Bolster, Matthew Burton, Min Kim, Andrew Bastawrous, Ari Ho-Foster, Hannah Chroston, Oathokwa Nkomazana

## Abstract

**Background:** Clinic non-attendance rates are high across the African continent. Emerging evidence suggests that phone-based reminder messages could make a small but important contributing to reducing non-attendance. We used behavioral economics principles to develop an SMS and voice reminder message to improve attendance rates in a school-based eye screening programme in Botswana.

**Methods:** We will test a new theory-informed SMS and voice reminder message in a national school-based eye screening programme in Botswana. The control will be the standard SMS message used to remind parents/guardians to bring their child for ophthalmic assessment. All messages will be sent three times. The primary outcome is attendance for ophthalmic assessment. We will use an automated adaptive approach, starting with a 1:1:1:1 allocation ratio. Patients will not be blinded,

**Discussion:** As far as we are aware, only one other study has used behavioral economics to inform the development of reminder messages to be deployed in an African healthcare setting. Our study will will use an adaptive trial design, embedded in a national screening programme. Our approach can be used to trial other forms of reminder message in the future.

**Trial registration:** ISRCTN:96528723. Registered 5^th^ January 2022, https://doi.org/10.1186/ISRCTN96528723

**Administrative information:** Note: the numbers in curly brackets in this protocol refer to SPIRIT checklist item numbers. The order of the items has been modified to group similar items (see http://www.equator-network.org/reporting-guidelines/spirit-2013-statement-defining-standard-protocol-items-for-clinical-trials/).

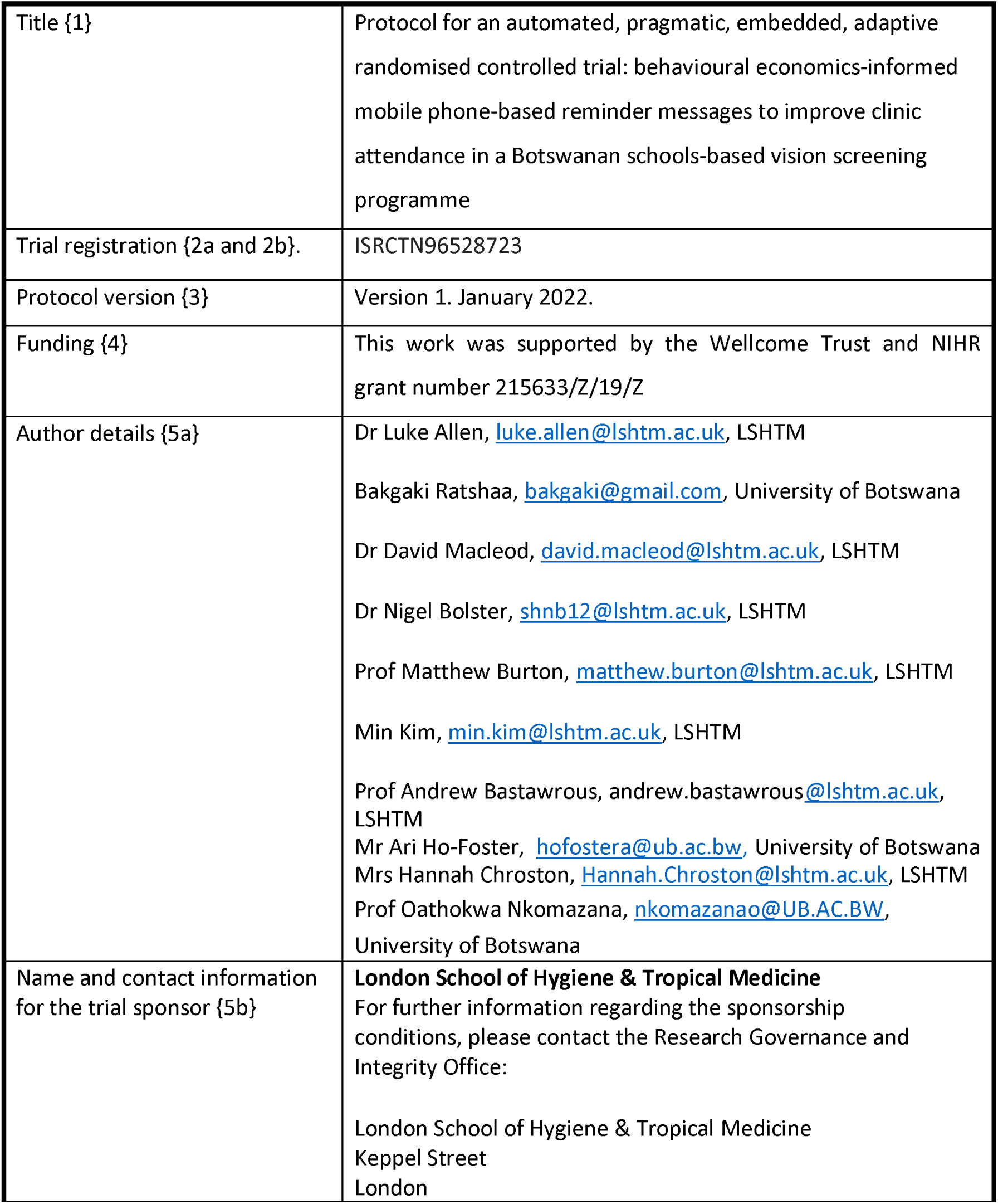

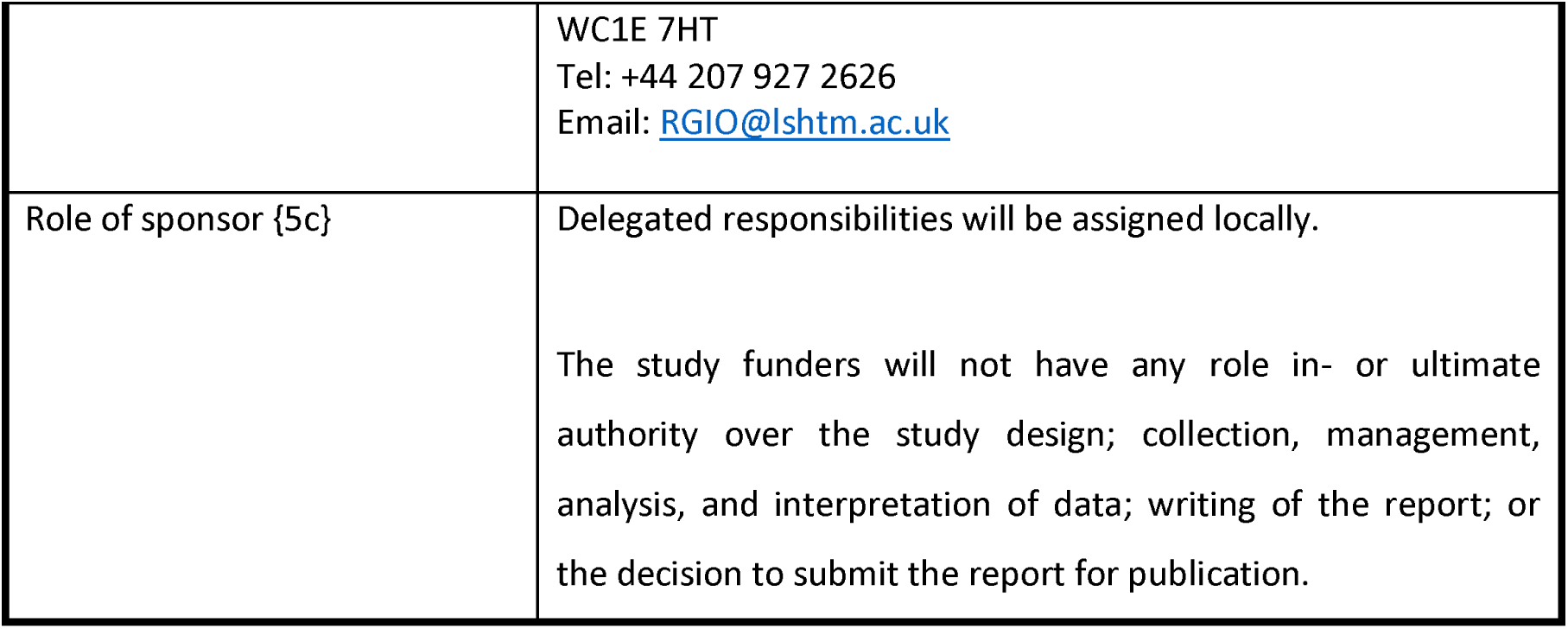

## Introduction

### Background and rationale {6a}

Many health programmes experience large mismatches between those identified with a clinical need and those who attend services. A recent international systematic review of ‘no-show’ appointments across all medical specialities in primary and secondary care estimated that 23% of clinic appointments are not attended, with the highest rate observed in the African continent (43%).^1^ Complex supply and demand factors govern access to health services,^2^ and systematically marginalised populations are often the least likely to receive care.^3,4^

As mobile phone penetration has risen, there has been increasing interest in the use of phone-based reminder messages to reduce these missed appointments. Systematic reviews from 2011,^5^ 2013,^6^ 2016,^7^ 2018,^8^ and 2019^9^ have found that SMS and voice-message reminders can improve clinic attendance by 50-100% depending on service, population and setting. In Linde and colleague’s systematic review of African RCTs, their pooled analysis found that SMS reminders doubled appointment attendance compared with no SMS (OR: 2·03; 95%CI: 1·40 to 2·95; I^2^ = 85%).^9^ Robotham and colleagues’ 2016 review found that “two or more notifications increased attendance by as much as 19% over and above sending one notification, and voice notifications may offer slight improvements over text notifications for increasing attendance.”^7^

SMS and voice messages function as behavioural ‘brief interventions’, and a number of studies have used behavioural economics principles to guide the wording of these messages in order to optimise their impact.^10,11^ Senderey and colleagues used a set of established cognitive biases to develop the content of eight different clinic reminder messages.^12^ Huf and colleagues used a similar approach in developing four different messages to boost clinic attendance in the UK^13^ Whilst both studies showed improvements in clinic attendance rates, neither provided rationale for why these specific biases were selected. In Linde and colleagues’ 2019 systematic review of 31 African RCTs using phone-based messages^9^ only one study reported using behavioral theories to develop the content of an SMS reminder message: Erwin and colleagues successfully boosted cervical cancer screening rates among women in Tanzania,^14^ basing their SMS reminder content on the Health Belief Model.^15^ They also reported using a motivational tone – found to be more effective than an informational tone^16^ – and pre-tested the SMS content validity and cultural sensitivity of the message with programme staff and laypeople.

One area that currently experiences very high rates of missed appointments – with substantial societal and economic costs – is community-based vision screening. Approximately 1.1 billion people (over 10% of the global population) currently live with a form of easily correctable visual impairment.^17,18^ Two very cheap and simple interventions - spectacles and cataract operations – could eliminate over 90% of all visual impairment worldwide.^17^ Provision of these services has risen exponentially in recent decades, however effective coverage rates are disappointingly low and exhibit marked socioeconomic gradients at the international and intra-national levels.^17^ Women and marginalised groups bear a disproportionate burden of visual impairment, and often face structural social barriers that prevent them from accessing care – as noted in the recent UN Resolution on Vision.^19^

Recognising the human, social, and economic drag exerted by cataracts and uncorrected refractive error, many LMIC governments are ramping up their vision screening programmes. Donor funding is rising in tandem, partly driven by the advent of phone-based screening platforms like Peek Acuity^20,21^ that have made it possible to rapidly screen entire regions with very low resource requirements.

Screening programmes based on the Peek digital platform are currently operating in seven low and lower-middle income countries (LMICs). The Botswanan Ministry of Health and Wellbeing (MoHw) is using Peek to screen all school children in government schools over three years beginning in 2022.^22^ The Peek platform records basic sociodemographic data, visual acuity, referral status, and attendance status for each child. Every time a child is referred a series of three SMS messages are sent to the mobile phone number registered by their parent/guardian (see Box 2). The current SMS message was not developed with reference to behavioural economics principles. According to internal data from Peek screening programmes in other countries, attendance rates are currently around 50% i.e., only half of those identified as needing ophthalmic assessment present to services.

We set out to develop two behavioural economics-informed reminder messages; an SMS and a pre-recorded voice message to be used in the new MoHw Botswanan schools-based vision screening programme, and tested using an embedded, pragmatic, adaptive RCT design.

### Objectives {7}

Our objectives are to test a behavioural economics-informed SMS reminder message and a pre-recorded voice message that will be sent to the parents/guardians of school children who have been identified as having a correctable visual impairment and referred to refractive services. We hypothesise that these messages will be associated with a higher attendance rate than the current standard SMS reminder that is sent to all referred patients’ parents/guardians.

### Trial design {8}

This is an automated, adaptive, parallel, four-arm, embedded, pragmatic RCT with an initial 1:1:1:1 allocation ratio. We will use a Bayesian adaptive trial algorithm to perform adaptive allocation as the trial progresses.

## Methods: Participants, interventions and outcomes

### Study setting {9}

Botswana schools-based vision screening programme.

### Eligibility criteria {10}

Reminder messages will be sent to the registered mobile phone numbers of parents/guardians of children who test positive at triage and are referred on for refractive services in the ‘Pono Yame’ MoHw/Peek Vision school screening programme in 2022. Provision of a mobile number is a pre-condition of entry into the screening programme, although parents/guardians are able to supply the number of a friend or relative.

Reminders will be sent in English and Setswana; spoken by >96% of the local population. The screening programme routinely collects data on preferred language and reminders will be sent in the preferred tongue. Those who list any language other than English of Setswana will receive a reminder in the dominant language (Setswana). We will perform a secondary-analysis that excludes these participants, but they will be included in the primary analysis.

### Who will take informed consent? {26a}

The interventions represent minor modifications to existing routine processes and present negligible risk to participants. Obtaining consent would introduce burdens to the participant that are greater than the intervention itself. As such, we will not seek informed consent. This approach has been approved by the LSHTM ethics committee and follows the precedent set by three previous RCTs testing SMS reminder messages.^12,13,23^

### Additional consent provisions for collection and use of participant data and biological specimens {26b}

All parents/guardians must provide consent for their children to partake in Peek Vision screening programmes. They are also routinely asked to consent to the use of their children’s sociodemographic data for research and sharing purposes. Care will not be compromised in any way for those participants who do not provide consent.

### Interventions

#### Explanation for the choice of comparators {6b}

The standard SMS message presented in Box 2 is routinely sent to the registered mobile phone of parents/guardians of children referred on for refractive services in all Peek programmes. This is the control arm.

#### Intervention description {11a}

##### Process of developing the intervention SMS and voice reminder messages

We aimed to use an established framework to identify a theory-informed set of behaviour change principles to guide the development of our reminder messages. We elected to use Dolan and colleagues’ *MINDSPACE* framework,^24^ developed in conjunction with the Institute for Government. This framework brings together insights from behavioral economics research that can be used to develop brief healthcare interventions (Table 1). The framework has been endorsed by the Behavioural Science and Public Health Network,^25^ the London School of Economics Behavioral Economics Playbook for behavior change,^10^ and the Health Foundation in their guidance on behavioral insights in health care.^11^

**Table 1:** The MINDSPACE framework and application for phone-based reminder messages.

| The nine principles | Application |
| --- | --- |
| Messenger | People are heavily influenced by the authority and credibility of the person sending the message, so the reminder messages should be signed-off by a trusted official/professional. |
| Incentives | People are more sensitive to losses than gains, so the reminders should frame non-attendance as a loss. |
| Norms | People want to fit in and are strongly influenced by the actions of others, so the reminders should signal that attendance is the norm. |
| Defaults | People tend to 'go with the flow' and use pre-set options, so attendance should be the default option in the reminders. |
| Salience | People are drawn to things that are novel and appear relevant to them, so the reminders should be personalised and stress the novelty of the opportunity. |
| Priming | Peoples' decisions are commonly influenced by subconscious cues in their environment. We cannot influence this via phone-message. |
| Affect | Peoples' decisions are often based on emotional associations rather than facts, so the reminders should seek to make emotive arguments for attendance. |
| Commitments | People seek to be consistent with public promises and reciprocate acts, so the reminders should aim to elicit a commitment to attend and stress the social expectation of attendance. |
| Ego | People act in ways that support the impression of a positive self-image, so the reminders should reinforce the message that attendance is consistent with recipients' positive self-perceptions. |

Whilst SMS and voice messages can include messenger, incentives, norms, defaults, salience, emotional appeals, commitments, and ego – they are less able to ‘prime’ recipients using subconscious cues. In addition to these principles, we also looked to the specific guidance on sending effective phone messages to reduce clinic non-attendance produced by Public Health England in 2020, based on their review of the international literature.^26^ Their key messages are summarised below:

- Messages should be clear, brief and well-formatted, with essential information only.
- Use line breaks to make the message easier to read.
- Personalise the text messages to include the recipient’s name if local systems allow.
- Keep messages to 480 characters (3 standard text messages) in length.
- Include the date, time and location of the appointment, as well as any special instructions, and contact phone number (if different to the number the text message is sent from).
- Write out the day of the week and the month in dates. For example, ‘Monday 23 March’.
- Make it easy to reschedule the appointment.
- Evidence suggests that GP endorsement can encourage people to take screening more seriously.

One researcher (LA) drafted an initial SMS that included all of the eight relevant MINDSPACE behavioral economics elements and adhered to PHE guidance (Figure 1). The names ‘Tebogo’ and ‘Dr Dineo’,the date, and the phone number are all made up. We convened a workshop to refine the SMS and develop a pre-recorded voice message with an African economist and representatives from the University of Botswana, and Peek Vision’s Botswana office.

**Figure 1:**
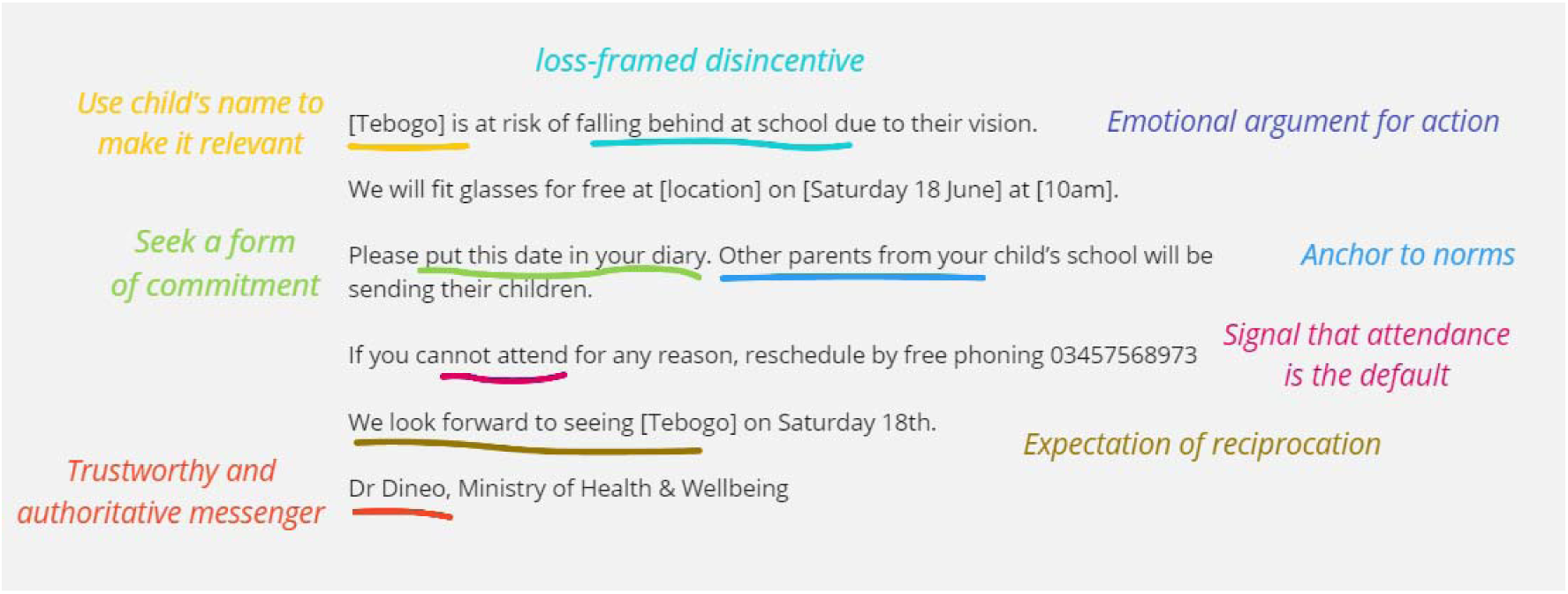
First draft SMS reminder.

##### Refinement process

The working group felt that the wording needed to be simplified so that parents/guardians with low levels of education would understand the messaging. The first sentence and the concept of ‘falling behind’ were felt to be too complex. We broke this sentence into two parts and simplified the language. The Peek team noted that refractive error is not the only problem that is managed, so we changed the wording around glasses to ‘eye problems’ to be more inclusive. The Botswanans in the group felt that we should drop the request to put the date in a diary as this may not be standard practice in many settings. We discussed whether the wording should request that participants make a commitment by responding to the SMS with the word ‘yes’, however this would incur costs on their behalf. Our economist – who also reviews ethics applications – felt that mentioning the behaviour of other parents may be felt to represent coercion rather than persuasion. We agreed on a compromise by stating that the screening service will be provided ‘for all children’. The group felt that ‘toll free’ was better wording than ‘free phoning’ and agreed that the default – and simplest option – should be to bring the child to the appointment. The group agreed to retain the final sentence in order to exert emotional reciprocity. It was agreed that a doctor would be the most appropriate authoritative messenger – rather than a professor or any other profession - and that the MoH represents the most authoritative and respected affiliate institution. We agreed to use the same wording for the voice message (Box 1).

###### Box 1

**Second draft following the workshop**

Hello, my name is Dr Dineo and I am calling from the Ministry of Health.

Your child recently had their eyes checked at school by the Pono Yame screening programme.

Unfortunately, your child was found to have an eye problem. If this is not corrected, it could affect their schoolwork.

Our medical team will be at your child’s school on [date/time]. Please bring your child to get a free medical assessment.

If you cannot come on that day, please call us to reschedule; We will text you a toll-free number that you can call.

If you can attend, there is nothing you need to do. Just bring your child on the day. We are offering this free assessment to all children with eye problems.

We look forward to seeing you and your child on [date/time]

Many thanks, Dr Dineo.

The message was then sent round for review from the wider research team. Wording revisions were suggested and agreed by consensus. Both messages were then translated into Setswana and back-translated into English, and checked by team members fluent in English and Setswana.

The final draft text was then tested with three Batswana laypeople (two teachers and a parent of a child with spectacles) using a qualitative ‘think aloud’ approach.[charters 2003] Each person was asked to read the messages and verbalise their thoughts. They were then asked to provide specific feedback to clarify and improve the wording. They recommended merging two sentences into one:

> **Original:** “We will do a free medical check-up for [name of a child]. We are doing this for all children with eye problems in the school.”
>
> **Recommended wording:** “We will be doing a free medical check-up for all the children with eye problems in the school.”

This feedback was incorporated into the final revision of the SMS and voice reminder messages by the original workgroup. The final intervention messages were translated and back-translated. The Setswana version was checked by Setswana speakers and the English back-translation was checked by two bilingual people. The final messages were approved by the research team and are presented in Box 1, alongside the standard (control) SMS. We note that PHE recommend aiming to fit the entire message into three 160 character SMS messages, however our message spills into four.

###### Box 2

**Control and intervention reminder messages**

###### Setswana

[name], [child’s name] o ne a tlhatlhobiwa kwa sekolong, a fitlhelwa a na le mathata a matlho. Tswe tswe mo tlise ko [location] go bona ba bongaka jwa matlho.

###### English

Dear [name], [child’s name] was examined at school and found to have an eye problem. Kindly report to [location] to see an eye specialist.

###### Setswana

Go motsadi:

Re lemogile ngwana wa gago [leina la ngwana] fa ana le bothata jwa matlho. Se, se ka ama tiro ya gagwe ya sekolo. Tswee-tswee, tsisa {leina la ngwana} ko sekolong ka (letsatsi le nako)

(Leina la ngwana) o tla tlhatlhobiwa matlho a sa duele

Se, se direlwa ngwana mongwe le mongwe mo sekolong yoo nang le bothata jwa matlho

Fa, o ka se kgone go tla, o ka rulaganya letsatsi le sele ka go leletsa mogala wa mahala wa (xxxxxxxxx).

Re ka leboga go le bona ka [letsatsi le nako]

###### English

Dear parent, we have found that your child [child’s name] has an eye problem. This may affect [his / her] schoolwork.

Please bring [child’s name] to [his/her] school at [time], [day, date]. We will be doing a free medical check-up for all the children with eye problems in the school.

If your child cannot come for any reason, please call us for free on number [xxxxxxxxxxxx]. We look forward to seeing [child’s name] on [day and time].

Many thanks, Dr [name], Ministry of Health

###### Setswana

Dumelang: Ke bidiwa ngaka Dineo, go tswa ko lephateng la botsogo. Ngwana wa gago [leina la ngwna] o tlhatlhobilwe matlho mo bogaufing, mme a fitlhelwa a na le bothata jwa matlho. Fa a ka seka a alafiwa, go ka ama tiro ya gagwe ya sekolo. Setlhopha sa rona sa botsogo, se tlaa bo se le ko sekolong sa ga [leina la ngwana] ka [letsatsi le nako].

Tswee.tswee tsisa ngwana wa gago go tlhatlhobiwa go sena dituelo. Se, se direlwa ngwana mongwe le mongwe yoo nang le bothata jwa matlho. Fa o ka seka wa kgona, tswee-tswee ikgolaganye le rona ko mogaleng wa [TBD xxxxxx] go re neela letsatsi lesele. Kea leboga [Leina la ngaka]

###### English

“Hello, my name is Dr [name] from the Ministry of Health.

Your child [child’s name] recently had [his/her] eyes checked at school and was found to have an eye problem. If this is not corrected, it could affect their schoolwork. Our medical team will be at your child’s school on [date/time]. Please bring your child to get a free medical assessment. This is offered to all children with eye problems.

If you cannot come on that day, please call us to reschedule on (XXXXXX). We will send you this number in a text message.

We look forward to seeing you and your child on [date/time] Many thanks, Dr [name].

We will use four arms as outlined below. Each SMS will be sent three times; on the day of referral, on the day before the appointment, and on the day of the appointment. To control for any effect the timing of the intervention might have, all SMS will be sent at the same time (6pm for the first two reminders, and 8am for the final reminder).

- Arm 1 (Control): Standard SMS reminder messages.
- Arm 2: New SMS reminder messages.
- Arm 3: Standard SMS reminder messages plus the pre-recorded voice reminder
- Arm 4: New SMS reminder messages plus the pre-recorded voice reminder

### Criteria for discontinuing or modifying allocated interventions {11b}

Due to the low-risk nature of the interventions, there will not be any formal option to discontinue or modify the reminder messages.

### Strategies to improve adherence to interventions {11c}

There are no relevant strategies to improve adherence. This is a pragmatic intention-to-treat study, and we will not collect data on whether messages were actually read or listened to by the intended recipients.

### Relevant concomitant care permitted or prohibited during the trial {11d}

No other reminder messages will be sent from the Peek platform during the trial.

### Provisions for post-trial care {30}

As this is a negligible risk trial, no provisions will be made for post-trial care.

### Outcomes {12}

All children who are screened and found to need further refractive assessment (e.g. for the fitting of glasses) will be given an appointment, 1-2 weeks later, at a specified time and location. The primary outcome is attendance at this pre-specified appointment within 3 weeks of the appointment date (yes/no). The Peek software retains a record of every referred child. When children attend for these appointments they are checked-in using Peek software. This automatically updates their attendance status. Attendance data will be automatically reviewed by an algorithm 24 hours. The great advantage of the Peek-based screening programme is that is a closed data system with complete, unified data records for every person screened, their referral status, and their attendance status. No additional data collection activities are required.

> **Primary outcome:** Attendance at refractive clinic within 3 weeks of invited date. This is a binary outcome measure (yes/no). We will compare mean outcome rates between arms.
>
> **Secondary outcome:** Days elapsed between referral and attendance. We will compare mean number of days elapsed between each arm.
>
> **Subgroup analyses:** Attendance by age, sex, urban/rural residence, distance to clinic, ethnicity, religion, language, household composition, migrant status, parental occupation, housing, assets and income.

### Participant timeline {13}

This automated adaptive trial will run continually for three months, recruiting participants until sufficient evidence has been gathered to reject the null (by triggering a stopping rule), or the trial enrols 3,000 participants – whichever comes first. Enrolment is planned to commence in Q2 2022. Poorly performing arms will be dropped but no new arms will be added.

**Figure 1:**
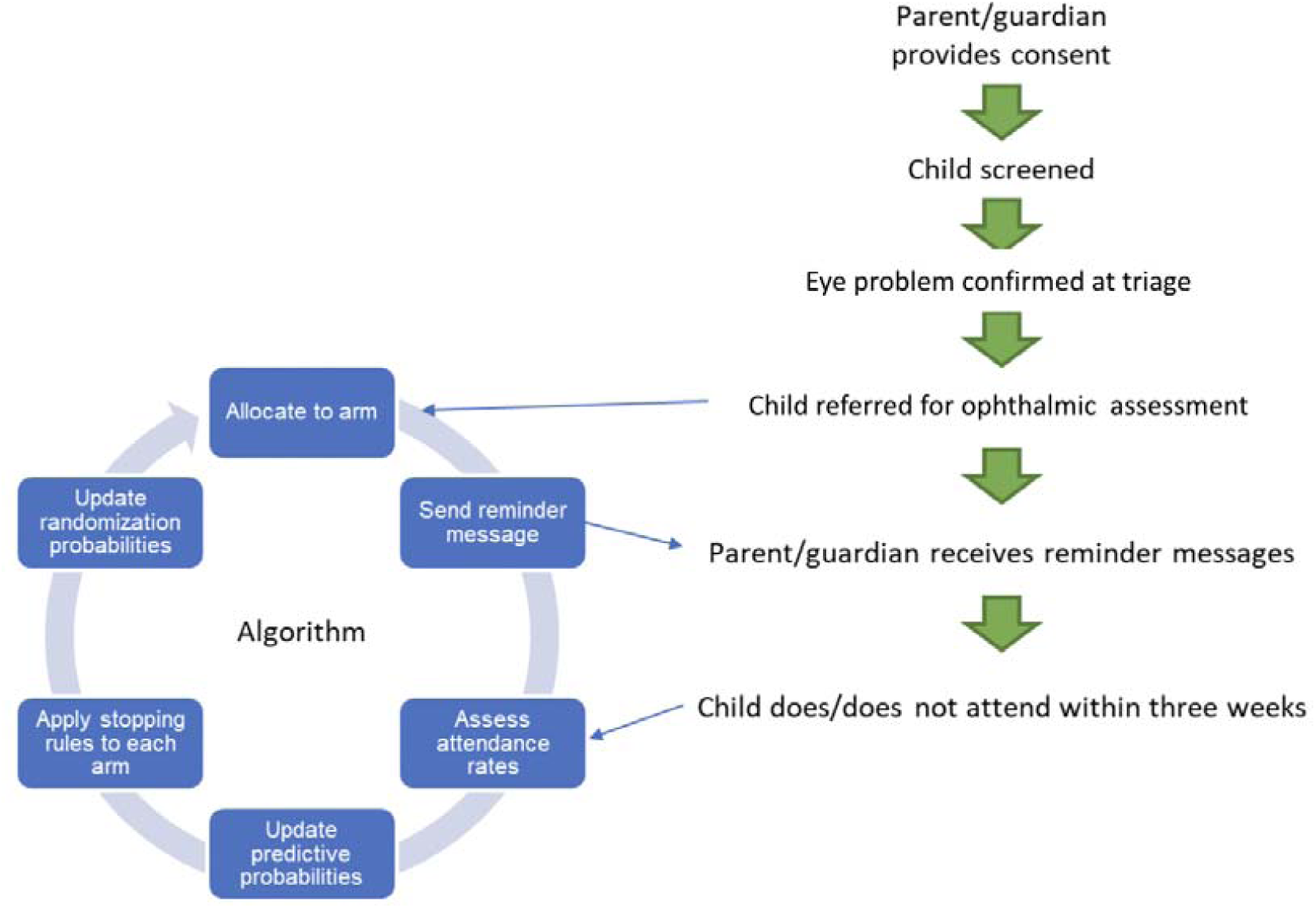
Interaction between patient flow and the adaptive trial algorithm.

### Sample size {14}

Approximately 6,500 children will be screened every month. Based on previous programmes, we expect approximately 1,000 of these children to be identified as requiring referral. All of these children’s parents/guardians would receive the standard three SMS reminders in a standard programme.

Using a traditional statistical analysis, we would perform three comparisons versus the control arm, so would use a Bonferroni adjusted alpha of 0.0167. Assuming 50% attendance on the expected day in the control arm we would need 667 referred children per group to have 90% power to detect an improvement to 60% attendance in the intervention arms (approximately 2700 in total). To detect an improvement to 55% we would require a sample size of 2,692 per group (approximately 10,700 in total). The disadvantage of the traditional approach to sample size calculations is that the assumptions are often incorrect, leading to over-powering which needlessly exposes participants to ineffective arms, or underpowering which wastes resources.

The adaptive allocation method that we are using does not depend on assumptions and does not use a pre-specified a sample size. Instead, the study will run until one of three criteria are met:

- There is a >95% probability that one arm is best.
- There is a >95% probability that the difference between the arms remaining in the study is <1%.
- 3,000 participants have been enrolled.

This approach is almost perfectly efficient, as exactly the right number of participants are enrolled to answer the research question. Depending on the effect of the interventions, one of the stopping criteria might be met after 100 participants, however it could also take 100,000 before reaching a definitive conclusion. We will set a ceiling on the number of total participants of 3,000 for this current study due to resource constraints.

As we anticipate that 1,000 children will be referred every month, our recruitment should last no longer than approximately three months (but potentially shorter if one of the first two criteria are met).

### Recruitment {15}

Every referred child’s data will be included in the primary analysis. Subgroup analyses will only be permitted for children whose parents have consented for their sociodemographic data to be used for research purposes. This is a separate consenting process is led by Peek.

### Assignment of interventions: allocation

#### Sequence generation, concealment, and implementation {16a, 16b, 16c}

Participants will initially be randomly allocated into four arms using computer-generated blocks of 12. As allocation and intervention delivery (sending SMS messages) is fully automated, there is no need for any of the human investigators to know participant allocation status. Once the first participants attend refractive services the algorithm will begin adjusting the allocation ratio to favour the best-performing arms. There is no need for the investigators to see allocation status at this stage either. The data safety monitoring committee will be fully unmasked to allocation status and all outcome data and will have the power to stop the trial or suspend any arm.

### Assignment of interventions: Blinding

#### Who will be blinded {17a}

Trial participants will not be blinded. Programme implementers will check in participants when they attend clinic using Peek Capture. The software will automatuically record the date and the time elapsed since referral. The adaptive algorithm will analyse attendance rates between arms according to pre-defined rules. Screening programme staff and data analysts will be blinded to assignment status. A small team of unblinded human statisticians will tasked with monitoring the algorithm’s performance. They will double-check the algorithm’s working at regular intervals during the trial and will repeat the final analysis comparing each arm. They will have the power to stop the trial, but they will not influence allocation.

#### Procedure for unblinding if needed {17b}

There is no procedure for unblinding.

### Data collection and management

#### Plans for assessment and collection of outcomes {18a}

Referral status, attendance status, and days elapsed since referral will collected using the Peek Capture system on Android devices. Every time a participant is referred and every time they attend at clinic they are checked in using an android device operating Peek Capture software. Additional data on sociodemographic characteristics will be collected when participants initially present to the screening programme.

#### Plans to promote participant retention and complete follow-up {18b}

As the intervention is an SMS sent automatically by the programme, there is no scope from deviation. Similarly ‘loss to follow up’ is a primary outcome (non-attendance at appointment).

#### Data management {19}

Data will be collected by Peek’s implementing partners using Android devices through the Peek Capture application. Peek Capture enforces security controls that include strong device passcodes and native Android encryption. Data stored is time limited, the device syncs via an encrypted connection with a Peek managed server, the data is then deleted to minimise the risk of data stored on the device.

Data will be stored on a Peek managed server hosted in a Virtual Private Cloud (VPC) utilising the Amazon Web Services (AWS) Cloud. Each Peek powered programme is hosted on its own dedicated server and a VPC that will reside in the UK/EU ensuring all of the data privacy safeguards as governed under the GDPR. All data collected is securely stored in AWS data centers which are state of the art, utilising innovative architectural and engineering approaches. Routine manual data cleaning will be conducted periodically by Peek administrators. Internal software guardrails will pick up simple errors.

#### Confidentiality {27}

Peek routinely collects sociodemographic information from each child who is referred on for refractive services using the following domains: age, sex, location, ethnicity, marital status, religion, migrant/refugee status, occupation, education, food adequacy, housing, and asset ownership. This information will be held on a Peek managed server hosted in a Virtual Private Cloud (VPC) utilising the Amazon Web Services (AWS) Cloud. Peek also seeks consent to use this data for research purposes.

Sociodemographic data on participants who have provided consent will be shared with the statistical analysis team at LSHTM for subgroup analysis. All team members who will access these data will have undertaken information security training. We will use encrypted data transfer and avoid cloud services outside the EU. The aggregated Peek data that is shared with LSHTM project staff will not contain any names, however the data being shared may still permit the identification of individuals depending on the domains being shared and may therefore constitute pseudo-anonymised data. All data arising from this project will be stored securely for 10 years. Further information is provided in the data management plan (Appendix).

#### Plans for collection, laboratory evaluation and storage of biological specimens for genetic or molecular analysis in this trial/future use {33}

Not applicable. We will not be using biological specimens.

### Statistical methods

#### Statistical methods for primary and secondary outcomes {20a}

This study will use a Bayesian approach to identify the best arm. Every 24 hours the probability of each arm being the best arm overall will be estimated, using Monte-Carlo simulations combined with non-informative priors to get the posterior probability estimates. Each arm will have a probability of being best that of between 0 and 100%, and the sum of all four probabilities will equal 100%. These probabilities will be compared to the stopping rules as to whether the trial should stop or continue into the next day. If the trial is to continue, the proportion allocated to each arm for the next day will be updated to be proportional to the estimated probabilities.

#### Interim analyses {21b}

This is an automated adaptive trial. Our algorithm will review the attendance data every 24 hours and perform statistical testing. Three stopping rules will be applied during these daily interim analyses:

1. There is a >95% probability that one arm is best.
1. There is a >95% probability that the difference between the arms remaining in the study is <1%.
2. 3,000 participants have been enrolled.

If none of these rules have been satisfied, then the trial (i.e. enrolment) will continue. The Bayesian algorithm will adjust the allocation ratio based on the performance of each arm with respect to the updated posterior probability that each is associated with attendance.

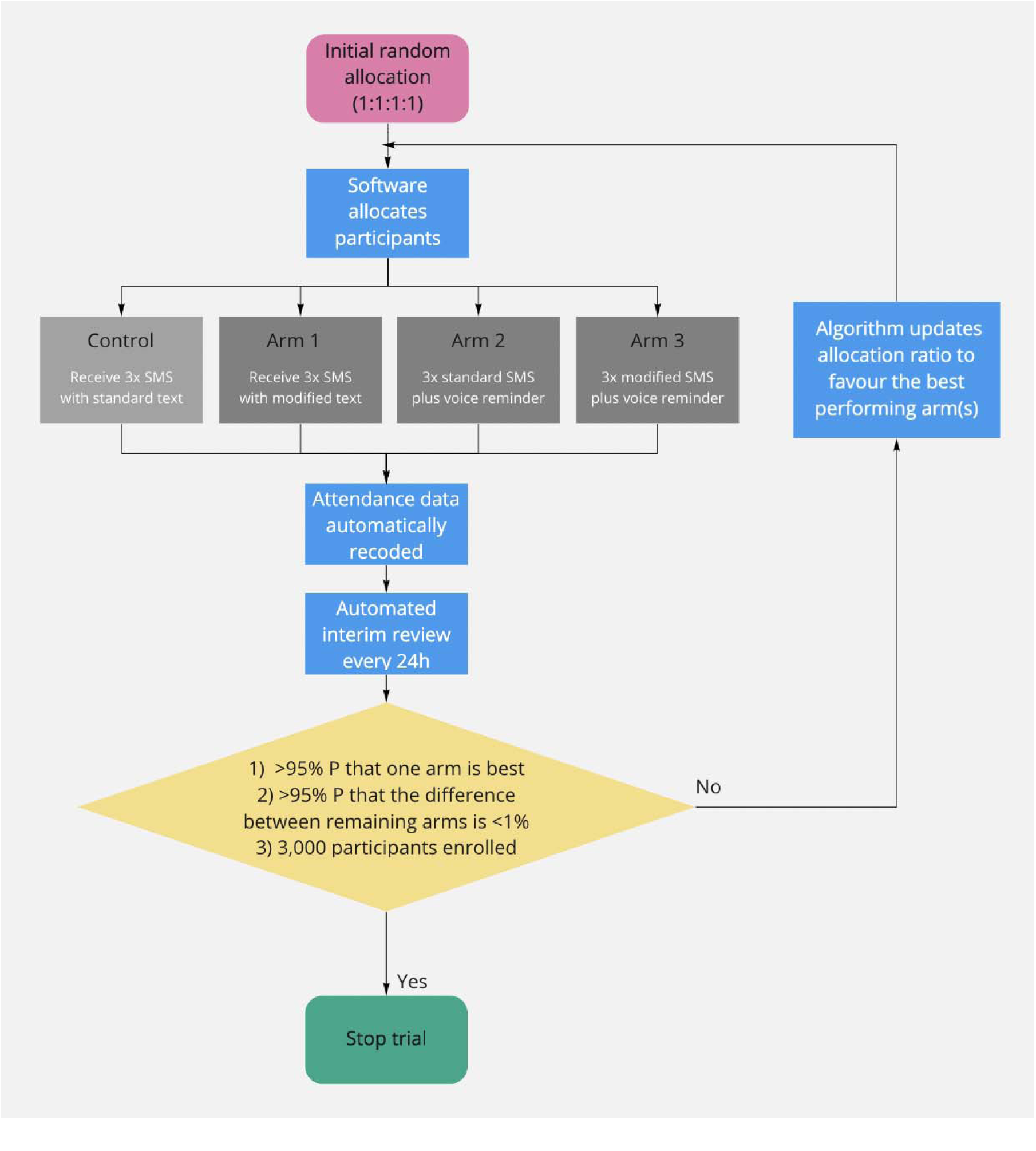

### Methods for additional analyses (e.g. subgroup analyses) {20b}

Once the trial is complete we will perform retrospective subgroup analyses to explore whether there is any evidence that attendance within each group was associated with sociodemographic variables. We use logistic regression to assess whether each sociodemographic variable is associated with attendance.

We will perform a secondary analysis that excludes participants whose preferred language is neither Setswana or English.

### Methods in analysis to handle protocol non-adherence and any statistical methods to handle missing data {20c}

The primary analysis only requires trial arm and the outcome (attendance) to be recorded. The trial arm should be recorded automatically as part of the Peek coding, and if it was missing it would be due to a bug in the coding. If this occurred there is no statistical method that could be used to recover that data so any records with trial arm missing would not be included in the updating of the probability that an arm is best. The outcome cannot be missing, as a participant is set as “not attended” until the point where they are updated as having “attended”, but if a record is missing in the database for some reason the record will be excluded from any analysis.

### Plans to give access to the full protocol, participant level-data and statistical code {31c}

The full protocol is available from the corresponding author. Statistical code will be made freely available online using GitHub. In line with the UK concordat on open research data (2016) anonymised participant-level data from this trial will be made available to bona fide research groups (evidenced via CVs and the involvement of a qualified statistician), and in line with the trial’s publicly available data sharing policy, following review and approval from the trial’s data monitoring committee. No reasonable request will be turned down, and the appropriate data will be made available within 1-month of receiving the request. There may be multiple levels of permission required in-country before data can be shared, including national ministry of health approval and local implementation partner approval.

### Oversight and monitoring

#### Composition of the coordinating centre and trial steering committee {5d}

##### Trial coordinating centre

- Dr Luke Allen, Co-PI and trial manager, LSHTM
- Hannah Chroston, lead administrator, LSHTM
- Bakgaki Ratshaa, trial coordinator, University of Botswana

#### Trial management group

- Dr Anrew Bastawrous, chief investigator
- Prof Oathokwa Nkomazana, co-PI
- Dr Luke Allen, co-PI
- Prof Matthew Burton, methods advisor
- Dr David Macleod, lead statistician
- Dr Nigel Bolster, Peek integration
- Min Kim, statistician
- Dr Michael Gichangi, methods advisor

#### Data management team

- Dr Luke Allen, co-PI
- Dr David Macleod, lead statistician
- Dr Nigel Bolster, Peek integration
- Min Kim, statistician

### Composition of the data monitoring committee, its role and reporting structure {21a}

An independent Data and Safety Monitoring Board (DSMB) will be appointed by the trial steering committee. The DSMB will have three members, all independent of the running of the trial with relevant clinical and epidemiological experience.

The DSMB will confirm their specific meeting arrangements. It is proposed that the DSMB would meet prior to the beginning of the trial (Q2 2022), one third of the way through, and at the end, to assess the safety of the trial procedures. The DSMB will agree the way it will monitor the data, what it requires from the investigators in this respect and will communicate this to the PIs. All data can be interrogated remotely in real-time.

The DSMB may visit the study coordination centre to assess data management, record keeping and other important activities. The DSMB will determine the manner in which it will monitor the data, what it requires from the investigators in this respect and will communicate this to the PIs.

### Adverse event reporting and harms {22}

#### DEFINITIONS

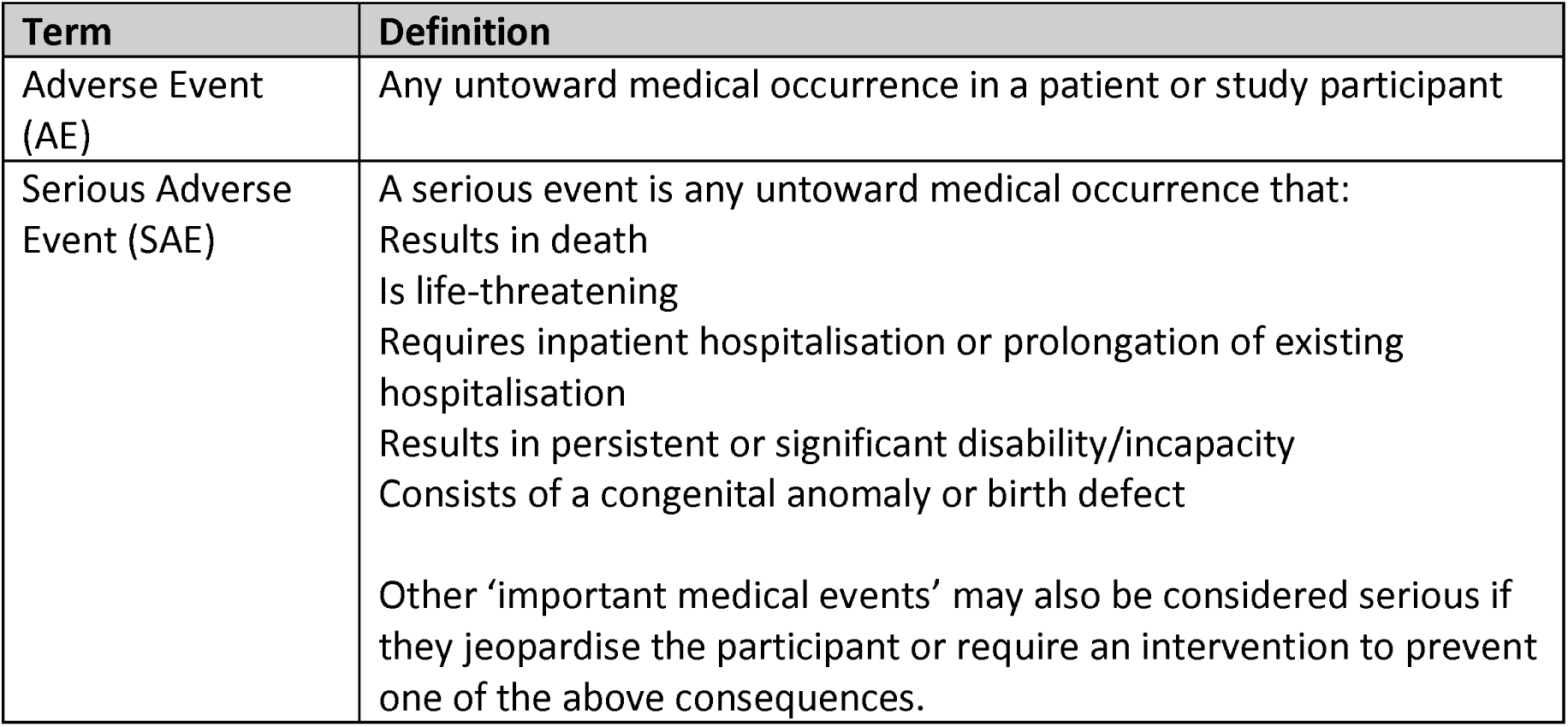

| Term | Definition |
| --- | --- |
| Adverse Event (AE) | Any untoward medical occurrence in a patient or study participant |
| Serious Adverse Event (SAE) | A serious event is any untoward medical occurrence that:<br>Results in death<br>Is life-threatening<br>Requires inpatient hospitalisation or prolongation of existing hospitalisation<br>Results in persistent or significant disability/incapacity<br>Consists of a congenital anomaly or birth defect<br>Other 'important medical events' may also be considered serious if they jeopardise the participant or require an intervention to prevent one of the above consequences. |

#### REPORTING PROCEDURES

All adverse events will be reported. Depending on the nature of the event the reporting procedures below will be followed. Any questions concerning adverse event reporting will be directed to the study coordination centre in the first instance. The flow chart below has been provided to aid the reporting of AEs.

### Responsible Personnel

#### Chief Investigator (CI)

- The CI has overall responsibility for the conduct of the study and the ongoing safety and evaluation of any IMPs being used in the trial.
- Promptly notifying all investigators, Institutional Review Board (IRB) or Independent Ethics Committee (IEC) and Competent Authorities (CAs) (i.e. LSHTM, EFMHACA, ORHB, FMOST, DSMB) of each concerned member state of any findings that may affect the health of the trial participants.
- Keeping detailed written reports of all AEs/ARs identified in the protocol as critical to the evaluation of safety within the agreed timeframes specified in the protocol.
- Accurate production and submission of the Development Safety Update Reports (DSURs) and progress reports to CAs and IRB/IECs.
- Collate all AR/AEs/SAEs/SARs and report to the Sponsor annually.
- Ensure that the PIs report all SAEs/SUSARs immediately to the Sponsor and to the CAs, IRB/IECs and any other relevant parties within agreed timelines (
- Supplying the Sponsor and IRB/IEC with any supplementary information they request.

#### Principal Investigators (PI)

- The PIs have responsibility for the research performed at the local site, handling and management of investigational medical products, and informing the CI, Sponsor, Ethics, regulatory bodies and the trial coordinating team, of all adverse events that occur at their site
- Safety responsibilities:
- Ensure trial participant safety and the swift and adequate management of trial participants with any type of AE/AR as per the management protocol described below.
- Reporting all SAEs/SUSARs immediately to the Sponsor and to the CAs, IRB/IECs and any other relevant parties within agreed timelines (i.e. LSHTM, EFMHACA, ORHB, FMOST).
- Assessing each event for causality, severity and expectedness. (Note: a medical decision which must be made by the investigator directly involved with the care of the patient/participant experiencing the AE)
- Ensure adequate archiving of AE records and reports in the local trial office along with the trial master files.
- Collate all AR/AEs/SAEs/SARs biannually and present to the CI.
- Guide and supervise the field research team on accurate recording, reporting of all adverse events.

#### Field Research Team Members (Coordinators, Nurses, Examiners, Recorders)

- All field research team members are responsible for identifying, recording, and reporting any AE or AR to the PIs regardless of severity or causality.
- Assessing each event for causality, severity and expectedness. (Note: a medical decision which must be made by the investigator directly involved with the care of the patient/participant experiencing the AE).
- Ensure that the participant has received the necessary management. This includes advice/reassuring, referral, offering transport, paying for management, making follow-up visits
- Report to the PIs/Project manager AEs/ARs based on the specified timeline and file all AE/AR recorded forms in the trial master file.

#### Non-serious AEs

All non-serious AEs will be reported to the study coordination centre and recorded in a dedicated AE log within 72 hours. The entry must state the patient ID, date and time of AE, nature, and relation to the intervention, if any. The AE should also be reported to the data and safety monitoring committee within 72 hours. AE logs will be stored on a secure, password-protected file on a LSHTM computer.

#### Serious AEs

Serious Adverse Events (SAEs) will be reported to the PI and study coordination centre within 24 hours of the local site being made aware of the event. The PI will report the event to the data safety monitoring committee within 48 hours and include it in the study safety report.

An SAE form will be completed and submitted to the PA and study coordination centre with details of the nature of event, date of onset, severity, corrective therapies given, outcome and causality. All SAEs whether expected, suspected or unexpected will be reported to regulatory bodies and the trial DSMB within 48 hours of occurrence. The responsible investigator will assign the causality of the event. All investigators will be informed of all SAEs occurring throughout the study. If awaiting further details, a follow up SAE report should be submitted promptly upon receipt of any outstanding information.

Any events relating to a pre-existing condition or any planned hospitalisations for elective treatment of a pre-existing condition will not need to be reported as SAEs.

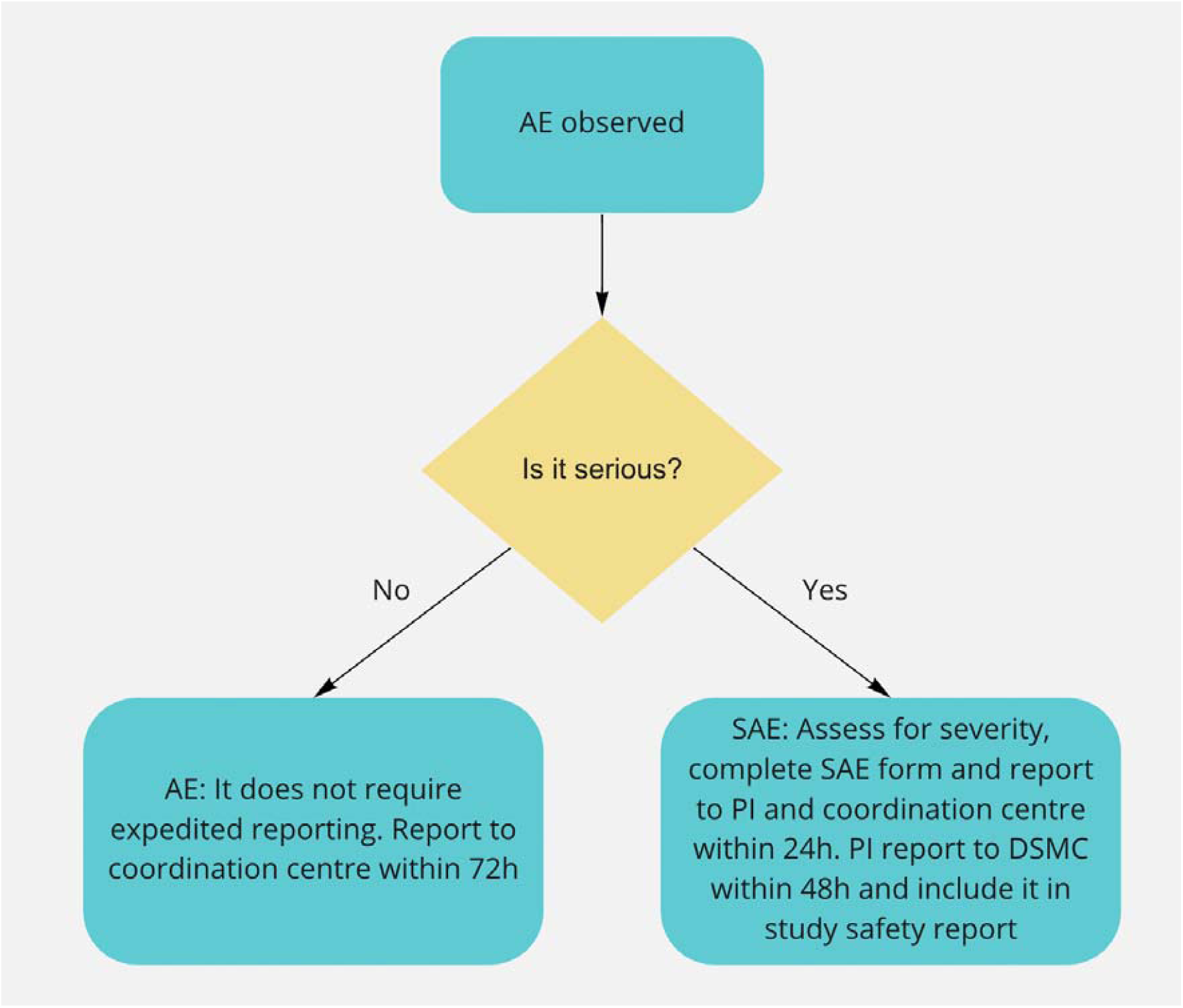

### Contact details for reporting SAEs

**Please send SAE forms to:** **or** **using the title ‘SAE’**

**Tel: +44 (0) 20 7958 8316 (Mon to Fri 09.00 – 17.00)**

**Tel**: +267 355 0000

#### Frequency and plans for auditing trial conduct {23}

The study may be subject audit by the London School of Hygiene & Tropical Medicine under their remit as sponsor, the Study Coordination Centre and other regulatory bodies to ensure adherence to Good Clinical Practice.

#### Plans for communicating important protocol amendments to relevant parties (e.g. trial participants, ethical committees) {25}

Any important protocol modifications will be reported to the co-investigators, research committees, the trial registry, and – where appropriate – journals and regulators via email.

### Dissemination plans {31a}

Scientific results will be published in Open Access in peer-reviewed journals and presented at relevant international conferences. All publications and presentations relating to the study will be authorised by the Trial Management Group. The first publication of the trial results will be in the name of the Trial Management Group members. Members of the Data and Safety Monitoring Board will be listed and contributors will be cited by name if published in a journal where this does not conflict with the journal’s policy. Authorship of any parallel studies initiated outside of the Trial Management Group will be according to the individuals involved in the project but must acknowledge the contribution of the Trial Management Group and the Trial Coordinating Centre.

## Discussion

This study is embedded in the national Pono Yame school-based vision screening programme. As such, any delays to the launch of the programme will delay the start of the trial. As far as we are aware, only one other study has used behavioral economics to inform the development of reminder messages to be deployed in an African healthcare setting. Our study will will use an adaptive trial design, embedded in a national screening programme. Our approach can be used to trial other forms of reminder message in the future, including tweaks to the messages that are sent and varying message content according to the demographic characteristics of the recipient.

### Trial status

This is protocol version 1.1 (10^th^ March 2022). Recruitment has not yet commenced but is planned for Q2 2022.

## Data Availability

No data are available

## Abbreviations

DSMC: Data and Safety Monitoring Committee
LSHTM: London School of Hygiene and Tropical Medicine
RCT: Randomised controlled trial
WHO: World Health Organisation

## Declarations

Authors’ contributions {31b}

- LA is co-PI. He led the protocol, co-designed the study, and drafted the manuscript.
- ON is co-PI. She conceived and co-designed the study, and revised and approved the manuscript.
- AB is the chief investigator; he conceived the study, led the initial funding proposal, and revised and approved the manuscript.
- DM is the lead statistician. He conceived and co-designed the study, revised and approved the manuscript, and developed the code.
- NB conceived and co-designed the study, revised and approved the manuscript, and integrated the code into the Peek software.
- MK conceived and co-designed the study, revised and approved the manuscript.
- MB conceived and co-designed the study, revised and approved the manuscript.
- BR developed the intervention and revised and approved the manuscript.
- AH-F co-designed the study and read and approved the final manuscript
- HC revised and approved the final manuscript.

## Availability of data and materials {29}

Any participants’ identifiable personal data collected following informed consent by the Trial Coordinating Centre will be stored in a secure Peek Vision server (See Data Management Plan for more details on the purpose and type of data collected, and the security of the Peek Vision server). Confidentiality protected in accordance with the Data Protection Act 2018 and the General Data Protection Act. Patient-level data will be pseudo-anonymised removing names and any other key identifiers before it is shared. Only the least amount of data will be shared, and where possible it will be fully anonymised and aggregated.

All published findings will be at anonymous aggregate subpopulation level. In line with the UK concordat on open research data (2016), anonymised data from this trial will be made available to bona fide research groups (evidenced via CVs and the involvement of a qualified statistician), and in line with the trial’s publicly available Data Management Plan (Appendix), following review and approval from the trial’s data monitoring committee. No reasonable request will be turned down, and the appropriate data will be made available within 1-month of receiving the request.

Peek Vision has a signed data sharing agreement with the Ministry of Botswana that governs the use of patient data collected and used in Peek Vision screening programmes. There may be multiple levels of permission required in-country before data can be shared, including national ministry of health approval and local implementation partner approval.

## Ethics approval and consent to participate {24}

The study will be conducted in accordance with the recommendations for physicians involved in research on human subjects adopted by the 18th World Medical Assembly, Helsinki 1964 and later revisions. The study will comply fully with the Botswana Data Protection Act of 2018.

In line with previous phone-based reminder message RCTs, this study tests a negligible risk intervention and will not seek informed consent from participants. The Study Coordination Centre has already recieved approval from the LSHTM Research Ethics Committee (Ref 26480). The Coordination Centre will also seek approval from the University of Botswana Research Ethics Board (Office of Research and Development) and the study will not commence until approval has been obtained

## Consent for publication {32}

Not applicable.

## Appendix: Data Management Plan

### Digital resources

Relevant details to mention: topics covered, type (e.g. survey), source (collected by self or others), format (e.g. STATA) and amount (e.g. 10 interviews). Draw attention to human or other data that require additional protection.

#### Data and Data Collection Process

The data will be collected in Peek powered Eye Health School and Community Programmes using Peek’s Capture application. During the Programmes initial screening process only basic and non personal identifying data is collected.

#### Data Fields Collected During the Initial Screening Process

- Age
- Gender
- Awareness (optional)
- Spectacle status
- Diabetes status (optional)
- Visual Acuity or pass/fail threshold
- Eye Condition

A representative sample of those screened will also be asked to provide sociodemographic data to enable us to monitor the equity performance of our programmes e.g. are certain ethnic groups more likely to be screened? The additional sociodemographic indicators are:

- Ethnicity
- Marital Status
- Religion
- Migrant/refugee status
- Occupation
- Education
- Food adequacy
- Housing (floor material)
- Asset ownership

Based on the visual acuity threshold set prior to screening the Peek Capture automatically informs the data collector whether the attendee may potentially need onward treatment. For those screened negative no further data is collected. Only for those screened positive is further information collected. This ensures data collection is kept to an absolute minimum maintaining privacy and ensuring compliance with data protection regulations.

For those screened positive additional information is collected, but the data is always minimised to ensure only the required data is collected at each stage of the service, as described below.

#### Data Fields Collected for Those Screened Positive at Triage

- Name
- Telephone number
- email address (optional)
- Visual Acuity

#### Further Data Fields Collected for Onward Treatment

- Eye Condition
- Prescription
- Diagnosis

We will also collect the sociodemographic details of all those referred on for further treatment:

- Ethnicity
- Marital Status
- Location (urban/rural)
- Religion
- Migrant/refugee status
- Occupation
- Education
- Food adequacy
- Housing (floor material)
- Asset ownership

#### Data Collection Tools

##### Data Storage

The data is stored on a Peek managed server hosted in a Virtual Private Cloud (VPC) utilising the Amazon Web Services (AWS) Cloud. Each Peek powered programme is hosted on it’s own dedicated server and a VPC that will reside in the UK/EU ensuring all of the data privacy safeguards as governed under the GDPR. All data collected is securely stored in AWS data centers which are state of the art, utilising innovative architectural and engineering approaches. More information, including a virtual tour, can be found by visiting the link below. https://aws.amazon.com/compliance/data-center/

### Hardware and software

#### Software

- Peek Capture - is an application that runs on Android devices that supports eye health screening and referral pathways to treatment
- Peek Admin - is a web based data platform application that is used to view the data collected by Peek Capture, it tracks the Programme progress, provides insights and helps ensure no one is left behind.
- STATA and R, and Excel will be used to analyse the data exported from Peek Admin

#### Hardware

- Peek servers are hosted on Amazon Elastic Compute cloud-based virtual machines running Amazon Linux.
- Android devices, locally managed by Peek’s implementing partners.

### Data-related activities

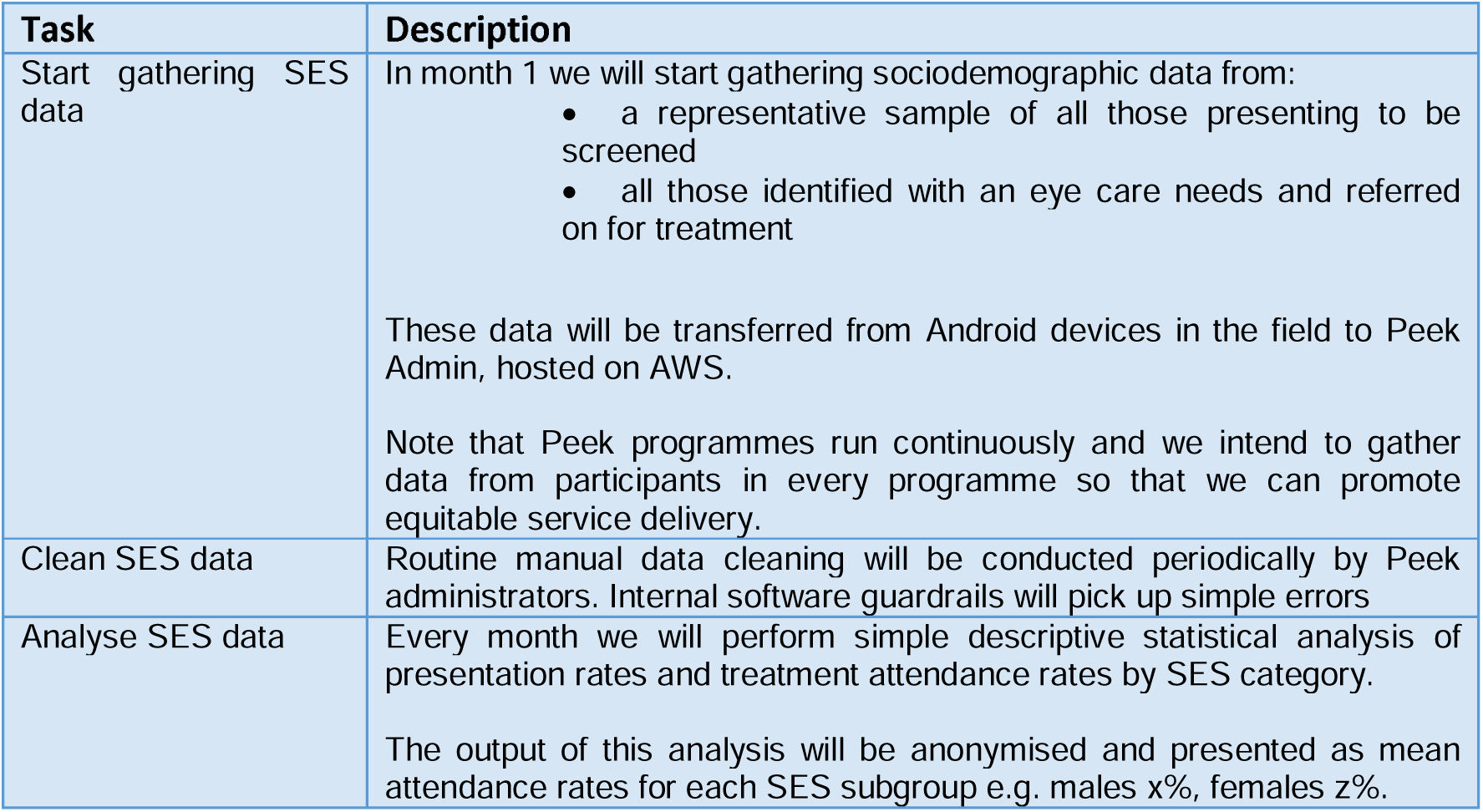

| Task | Description |
| --- | --- |
| Start gathering SES data | In month 1 we will start gathering sociodemographic data from:<br>• a representative sample of all those presenting to be screened<br>• all those identified with an eye care needs and referred on for treatment<br>These data will be transferred from Android devices in the field to Peek Admin, hosted on AWS.<br>Note that Peek programmes run continuously and we intend to gather data from participants in every programme so that we can promote equitable service delivery. |
| Clean SES data | Routine manual data cleaning will be conducted periodically by Peek administrators. Internal software guardrails will pick up simple errors |
| Analyse SES data | Every month we will perform simple descriptive statistical analysis of presentation rates and treatment attendance rates by SES category.<br>The output of this analysis will be anonymised and presented as mean attendance rates for each SES subgroup e.g. males x%, females z%. |

### Quality checks

Outline any quality checks to be performed before, during and after the above activities, e.g. to ensure data are captured correctly, remain accurate and complete, or ensure you avoid recognised problems. The UK Data Services offers guidance at http://ukdataservice.ac.uk/manage-data/format/quality.aspx.

- Errors are flagged at the point of data entry by software that only accepts pre-specified responses e.g. phone numbers must be comprised of a set string length of digits.
- The software has built-in logic steps
- We will institute training and supervision for all data collectors
- Application logging, audit trails and alerting direct administrators to given issues post-collection e.g. when SMS messages fail to be delivered
- Post-collection human data checking using the Peek Admin programme e.g. for ID disambiguation

### Ethical & legal issues

Local permissions for Peek powered eye health programmes are already in place. This is in the form of data processing agreements with Peek and the local MoH and/or local implementing partner. This provides a legal agreement between the parties that the data can be collected and processed. The proposed research will be authorised by the same parties to ensure full transparency and the data collection and processing will be managed under the same data processing agreement.

Informed consent is not required to run the phone-based reminder trial. LSHTM ethics board have approved this aopproach. We will not commence until the protocol has been approved by the University of Botswana ethics board, and approval from the government (HRU) board.

#### Consent processes used for other elements of the screening programme and embedded research

The Peek programme routinely collects sociodemographic data on all participants. This is independent of the current RCT. For this SES data collection we obtain written informed consent to collect, analyse, and publish anonymised aggregate participant data in peer-reviewed journals and online open-access data repositories. Individuals will not be identifiable. In line with UK guidance on risk-adapted approaches to obtaining informed consent, participants provide consent by ticking a box underneath the following statement:

*“I understand that my anonymous data may be shared with other researchers or online. I understand that I will not be identifiable from this information. I understand that my decision will not affect the care that I receive, and I am free to change my mind anytime I like*.*”*

Consent is obtained when participants initially present for screening. For screening programmes that include children (<18 years), we will seek consent from their parents/legal guardians using the following statement, sent home on a paper form along with the generic participant information leaflets:

*“I understand that my child’s anonymous data may be shared with other researchers or online. I understand that my child will not be identifiable from this information. I understand that my decision will not affect the care that my child receives, and I am free to change my mind anytime I like*.*”*

Approval will be sought from research ethics committees at LSHTM and each of the countries where screening takes place.

### Documentation

- Standard operating procedures and an overall study protocol will be developed in line with
- LSHTM research guidance to cover all aspects of the research project.
- Standardised online training modules have been delivered for programme implementing partners tasked with data collection in the field.
- Training will be delivered to all project staff to ensure that they understand the requirements and are able to follow the SOPs.
- We have a data compendium which describes the custom sociodemographic variables that we will collect in each country, available here.

### STORAGE AND SECURITY

#### Pre research data collection and storage in Peek powered eye health programmes

The data will be collected in Peek powered Eye Health School and Community Programmes using Peek’s Capture application. Data will be collected by Peek’s implementing partners using Android devices through the Peek Capture application. Peek Capture enforces security controls that include strong device passcodes and native Android encryption. Data stored is time limited, the device syncs via an encrypted connection with a Peek managed server, the data is then deleted to minimise the risk of data stored on the device.

The data is stored on a Peek managed server hosted in a Virtual Private Cloud (VPC) utilising the Amazon Web Services (AWS) Cloud. Throughout the eye health programme life cycle only approved implementation partners and Peek team members have access to programme data. Access is strictly controlled through the Peek Admin web based data platform application. This is used to view the data collected by Peek Capture, it tracks the Programme progress, provides insights and helps ensure no one is left behind. Peek Capture security:

- Peek Capture is installed on implementing partners managed Android devices
- Peek Capture enforces security controls that include strong device passcodes and native Android encryption.
- Data stored is time limited, the device syncs via an encrypted connection with a Peek managed server, the data is then deleted to minimise the risk of data stored on the device.

### Peek Admin security

- Strong passwords, minimum of 12 characters, password strength meter where only ‘strong’ is accepted, blacklist passwords are enforced to ensure easily guessed and passwords found in data breaches cannot be used.
- 2-Factor Authentication to protect user account security.
- User access permissions are controlled through account privileges, this controls scope of programme so access is restricted and limited to only what a user requires for their work, admin privileges are restricted to only those that require the access, account management and patient level reporting.
- Accounts disable automatically after 60 days of inactivity.
- User access reviews available for implementing partners to ensure leavers and inactive accounts are removed.

#### Peek Platform Data Security Assurance

Peek is an International Standardisation Organisation (ISO) 27001 certified organisation. ISO 27001 certification requires an annual audit by an accredited external auditing body who verify compliance with the industry best practice information security controls.

Peek servers hosted in a Virtual Private Cloud (VPC) utilising the Amazon Web Services (AWS) Cloud. Each Peek powered programme is hosted on it’s own dedicated server and a VPC that will reside in the UK/EU ensuring all of the data privacy safeguards as governed under the GDPR. All data collected is securely stored in AWS data centers which are state of the art, utilising innovative architectural and engineering approaches.

More information, including a virtual tour, can be found by visiting the link below:

https://aws.amazon.com/compliance/data-center/.

Annual penetration tests conducted by a 3rd party specialist security testing company. The purpose of the test is to verify whether robust security mechanisms are in place to prevent unauthorised users from accessing data and infrastructure. This penetration test includes:

- Identification of potential vulnerabilities occurring in the application and defining possible attack scenarios conducted with techniques typical for attacks on web applications;
- Simulated attacks from the perspective of an anonymous and standard user;
- Testing API endpoints from the perspective of an anonymous and standard user, including mechanisms such as user authentication, access control, and data validation;
- Security assessment of our infrastructure against the latest industry standard AWS CIS Foundations Benchmark.

The AWS Compliance Program provides further assurance and understanding of the robust controls in place to maintain security and compliance in the cloud. AWS regularly achieves third-party validation for thousands of global compliance requirements that are continuously monitored to meet security and compliance standards for the most sensitive data and privacy requirements. AWS supports more security standards and compliance certifications than any other offering, including PCI-DSS, HIPAA/HITECH, FedRAMP, GDPR, FIPS 140-2, and NIST 800-171, helping satisfy compliance requirements for virtually every regulatory agency around the globe. More information can be found by visiting https://aws.amazon.com/compliance/programs/.

#### Peek Platform Data Security Controls

##### Peek Servers

Peek servers hosted in a Virtual Private Cloud (VPC) utilising the Amazon Web Services (AWS) Cloud. Each Peek powered programme is hosted on it’s own dedicated server and a VPC that will reside in the UK/EU ensuring all of the data privacy safeguards as governed under the GDPR.

Server OS is Amazon Linux ustlising AWS AMIS to provide base images for our system drives and enhances security by focusing on two main security goals, limiting access and reducing software vulnerabilities. Security updates are applied automatically to test once a week and then rolled out a week later automatically to other environments

##### Docker

Peek server software runs in Docker containers. Docker shields application software from variations in platform and co-hosted software. It ensures that development, test and production environments run the same context as one another to ensure consistent, predictable behaviour. Peek servers also use docker swarm mode to achieve failsafe reliability and replication of Mongo databases.

##### Databases

Server data is stored in Mongo databases, a fast, scalable, json document database. Peek infrastructure uses a Mongo replica set across two hosts. There are two replicas each holding a full copy of the data and one arbiter. The arbiter is only used for the election of a new master if one of the nodes was to become unavailable. The Mongo database and journal are held on AWS Secure EBS volumes. This provides 256-bit AES encrypted using a key managed under the Amazon Key Management Service.

Amazon Key Management Service, allows us to create and manage cryptographic keys and securely control their use across a wide range of AWS services and within our applications. AWS KMS is a secure and resilient service that uses hardware security modules that have been validated under FIPS 140-2 to protect the encryption keys. AWS KMS also integrates with AWS CloudTrail providing us with secure logs of all key usage. Backups on S3 are also encrypted using keys managed by AWS Key Management Service.

##### Logging and Monitoring

Peek Server and Mongo Server logs and uploaded to AWS Cloudwatch for storage and monitoring. AWS Cloudwatch collects monitoring and operational data in the form of logs, metrics, and events and alerts us immediately of problems in any environment, both application and infrastructure.

##### Network Security

AWS Security groups are used to provide firewall-like network access control and allow inbound traffic on HTTP and HTTPS ports. Outbound traffic is permitted on any port. The SSH traffic is restricted to subnets associated with devops engineers and the deployment servers. TLS 1.2 is used to secure traffic between device or browser and server.

Operational access to the AWS console is protected with AWS IAM MFA which uses 2-Factor Authentication and ensures that access to AWS is restricted to users with knowledge of password and possession of a specific approved mobile device. Automated access to the AWS API uses AWS Roles with restricted privileges needed for housekeeping, logging and alarm maintenance. No user use is made of Access Keys to eliminate the vulnerabilities of file-system-based credentials.

##### Threat Detection

AWS Guard Duty is enabled, this provides a threat detection service that continuously monitors for malicious activity and unauthorised behaviour to protect access, workloads and data. The service utilises up-to-date threat intelligence feeds from AWS, CrowdStrike, and Proofpoint and continuously evolves through machine learning.

##### Backups

An Image is maintained of the Server Host using AWS AMI to ensure continuous availability.

A snapshot of the encrypted data volume, containing database and journal, is taken four times daily. Snapshots are retained for two weeks. Access to the snapshots is strictly controlled. Old backups are automatically deleted after 90 days. Backups are stored on AWS S3 storage, also encrypted providing 256-bit AES encryption. The backups are stored across AWS multiple availability zones, this ensures that the data resides in multiple data centres separated geographically and stored in AWS secure data centres.

Additionally a further backup is made off AWS. Off-AWS backups are replicated to Google Cloud daily via Google Transfer service to identically named buckets and files with a retention policy of 90 days.

##### Data Centres

All data collected is securely stored in AWS data centers which are state of the art, utilising innovative architectural and engineering approaches.

##### Disaster Recovery

A full disaster recovery test is performed at least annually to ensure servers, applications and databases can be fully recovered within 24 hours.

##### Export/data sharing for analysis

At the analysis stage pseudo-anonymised data will be exported in an encrypted zip file CSV file to LSHTM researchers to perform statistical testing. The zip file will be saved on the protected LSHTM server and only named project staff will be given access. Passwords will be sent separately. We will only ever export the minimum data required for the analyses.

### Labelling conventions

1. Keep file names short, meaningful and easily understandable to others.
2. Order the elements in a file name in the most appropriate way to retrieve the record.
3. Avoid unnecessary repetition and redundancy in file names and paths
4. Avoid obscure abbreviations and acronyms. Use agreed University abbreviations and codes where relevant.
5. Avoid vague, unhelpful terms such as “miscellaneous” or “general” or “my files”
6. Use capital letters to delimit words, as the preferred option, although underscores (_) or hyphens (-) may add clarity, they make the file name longer.
7. For numbers 0-9, always use a minimum of two digit numbers to ensure correct numerical order (e.g. 01, 02, 03 etc.)
8. Dates should always follow same format: YYYYMMDD e.g. 20170425
9. When including a personal name give the family name first followed by initials, with no comma in between e.g. SmithAB
10. Avoid using common words such as ‘draft’ or ‘letter’ at the start of file names unless doing so will make it easier to retrieve the record.
11. Use alphanumeric characters i.e. letters (A-Z) and numbers (0-9). Avoid using invalid characters in file names such as *? \ / : # % ~ { }
12. The file names of records relating to recurring events should include the date and a description of the event, except where the inclusion of these elements would be incompatible with rule 3.
13. The version number of a record should be indicated in its file name by the inclusion of ‘V’ followed by the version number (e.g. V01, V03 etc.). However versioning is enabled automatically in systems such as Office 365 and One Drive for Business, making it unnecessary to duplicate this information in the file name itself. e.g. 2021-11-19_Topic_Filename-variable01

### Measures to keep data safe and secure

|  |  |  |  |  |  |
| --- | --- | --- | --- | --- | --- |
| Only anonymised data will be used - personal, sensitive, or otherwise confidential data is not needed for the research |  | Store personal details in a separate secure location & link it via an identifier | Not required | Delete personal & confidential details at earliest opportunity (specify when below) | X |
| Use digital storage that require a username/password or other security feature | X | Physical security (such as locked cabinet or room) | X | Protect portable devices using security features, e.g. biometric |  |
| Encrypt storage devices | X | Encrypt during transfer | X | Avoid cloud services located outside EU | X |
| Take 'Information Security Awareness training' | X | Ensure backups are also held securely | X |  |  |
| Notes: | The aggregated Peek data that is shared with LSHTM project staff will not contain any names, however the data being shared may still permit the identification of individuals depending on the domains being shared and may therefore constitute pseudo-anonymised data.<br>We also note that there is not adequate shared secure storage space at LSHTM. We will have to use our personal H drives which is suboptimal for joint working and version control. |  |  |  |  |

### ARCHIVING & SHARING

- All data will be stored for 10 years.
- Internal confidential files will be retained on Peek’s secure servers.
- LSHTM analyses will be saved on encrypted and password-protected files on LSHTM SharePoint, with access restricted to the project team. Once the project is complete these files will be moved to a secure server.
- Data presented in publications (anonymised aggregate mean attendance rates for each SES subgroup) will be published on GitHub.

### When will the resources be made available?

|  |  |  |  |  |
| --- | --- | --- | --- | --- |
| During the research life |  | At the same time as findings are published in an academic journal | x | A set time after research end, e.g. 12 months. Specify below |
| Resources already available (provide details below) |  | On completion of my thesis |  | Other (provide details below) |
| Further information / Other |  |  |  |  |

### How will you make other researchers aware that the resources exist?

|  |  |  |  |
| --- | --- | --- | --- |
| Publish a metadata record describing the resources in a repository or other catalogue |  | Obtain a Digital Object Identifier (DOI) or other permanent ID |  |
| Cite resources in future research papers, e.g. in the data access statement or reference list | x | Cite resources in project reports | x |
| Publish a description for the project website |  | Write and publish a Data Paper |  |
| Add resources to a list of your academic outputs | x |  |  |
| Other measures / Further details |  |  |  |

### What steps will you take to ensure resources are easy to analyse and use in future research?

|  |  |  |  |
| --- | --- | --- | --- |
| Prepare a codebook or other documentation that provides an accurate description of content |  | Store resources in open file formats such as CSV, Rich Text, etc. See https://www.ukdataservice.ac.uk/manage-data/format/recommended-formats | x |
| Write a user guide that provides a high-level overview of research |  | Apply a standard licence that allows a broad range of uses (e.g. Creative Commons, Open Data Commons) |  |
| Designate a corresponding author / data custodian who will handle data-related questions | x | Use domain-specific standards that make it easy to import and analyse data |  |
| Other / Further information |  |  |  |

### If resources can be made available, but not openly, what conditions on access/use must be met?

E.g. data can be used for specific types of research only. Leave blank if not applicable.

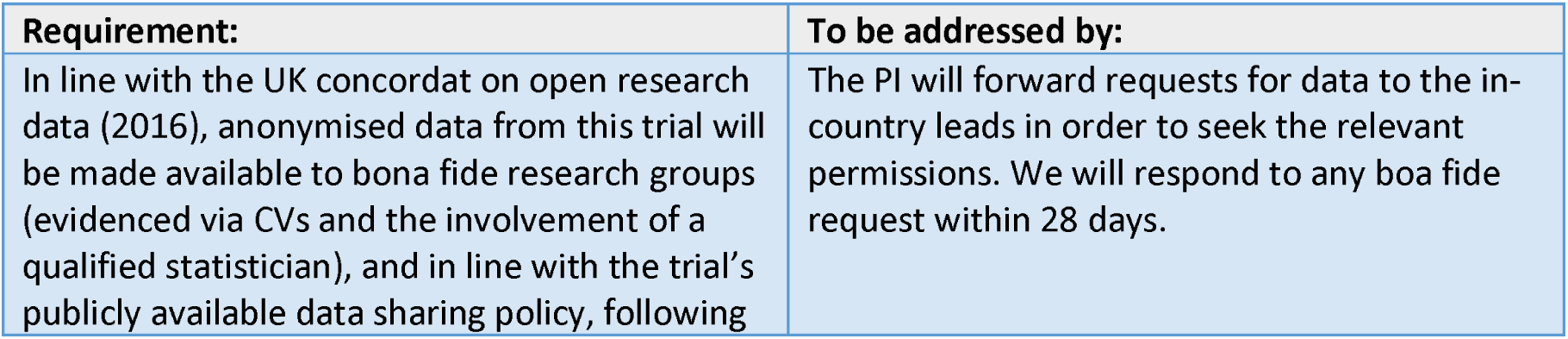
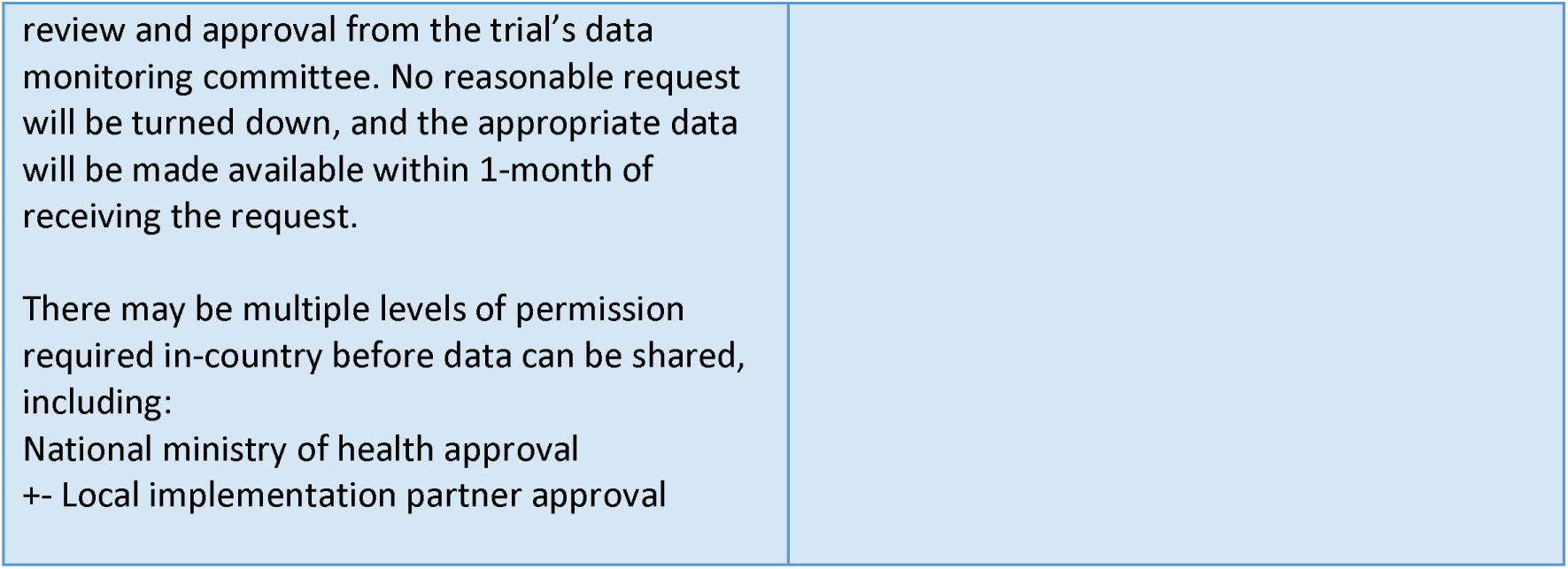

If resources can be made available, but not openly, what conditions on access/use must be met? E.g. data can be used for specific types of research only. Leave blank if not applicable.
| Requirement: | To be addressed by: |
| --- | --- |
| In line with the UK concordat on open research data (2016), anonymised data from this trial will be made available to bona fide research groups (evidenced via CVs and the involvement of a qualified statistician), and in line with the trial's publicly available data sharing policy, following | The PI will forward requests for data to the in-country leads in order to seek the relevant permissions. We will respond to any bona fide request within 28 days. |
| review and approval from the trial's data monitoring committee. No reasonable request will be turned down, and the appropriate data will be made available within 1-month of receiving the request.<br>There may be multiple levels of permission required in-country before data can be shared, including:<br>National ministry of health approval<br>+- Local implementation partner approval |  |

### RESOURCING

#### Primary data management challenges

With respect to costs of resources, we have adequate funding within the Wellcome project grant. The data is collected through active live Peek powered programmes where funding and resources is already provided for data collection and data security.

